# What is the quality of evidence informing vaccine clinical practice recommendations in Australia?

**DOI:** 10.1101/2025.03.05.25323463

**Authors:** Michael Dymock, Julie A Marsh, Kevin Murray, Thomas L Snelling

**Author notes:** Corresponding Author: Michael Dymock The Kids Research Institute Australia, Perth Children’s Hospital, 15 Hospital Avenue, Nedlands, 6009, Western Australia, Australia.

## Abstract

Policy and clinical practice guidelines are dependent on high quality evidence when making vaccine recommendations. A review of the quality of the evidence used to inform vaccine clinical practice guidelines could help guide researchers on how to improve the design of their clinical studies to produce evidence of greater value to decision-makers. In Australia, the Australian Technical Advisory Group on Immunisation (ATAGI) develops evidence-based vaccine clinical practice recommendations using the Grading of Recommendations Assessment, Development and Evaluation (GRADE) methodology, a transparent, systematic and methodical framework for developing and presenting summaries of evidence and its certainty.

We summarised the publicly available Australian GRADE assessments for the use of vaccines for prevention of cholera, diphtheria, tetanus and pertussis, human papillomavirus, influenza, meningococcal, pneumococcal, rabies and varicella zoster virus, including the certainty of evidence for each outcome (e.g., effectiveness, immunological or safety outcomes) and overall, in addition to the reasons for downgrade or upgrade of the certainty assessments. Across 25 research questions, 189 separate outcomes were assessed; of these 43 (22.8%), 38 (20.1%), 68 (36.0%) and 40 (21.2%) were classified as informed by *very low*, *low*, *moderate* and *high* certainty of evidence, respectively. Overall, 4 (16%), 10 (40%), 9 (36%) and 2 (8%) research questions across the disease areas had their overall certainty of evidence classified as *very low*, *low*, *moderate* and *high*, respectively. Certainty of evidence was downgraded for confounding, uncertainty in the effect estimation, and differences between the research questions asked by ATAGI and those answered in the studies.

There is an unmet need to improve the quality of evidence available to vaccine policy-makers. This could be achieved by improvements in the design of vaccine trials, particularly focusing on improving the precision of statistical estimates, inclusion of relevant subpopulations and ensuring trial endpoints are better aligned with the needs of policy-makers.

*Abbreviations:* ACIP, Advisory Committee on Immunization Practices; ATAGI, Australian Technical Advisory Group on Immunisation; COVID-19, coronavirus disease of 2019; DTP, Diphtheria, tetanus and pertussis; GRADE, Grading of Recommendations Assessment, Development and Evaluation; HPV, Human papillomavirus; JCVI, Joint Committee on Vaccination and Immunisation; NCIRS, National Centre for Immunisation Research and Surveillance; NHMRC, National Health and Medical Research Council; PCR, Polymerase chain reaction; PICO, Population, Intervention, Comparator and Outcomes; RCT, Randomised clinical trial; WHO, World Health Organization; Zoster, Varicella zoster virus; 13vPCV, 13-valent pneumococcal vaccine; 15vPCV, 15-valent pneumococcal vaccine; 20vPCV, 20-valent pneumococcal vaccine; 23vPPV, 23- valent pneumococcal polysaccharide vaccine.

## Introduction

Vaccine programs are a central strategy for the prevention of infectious diseases worldwide and are undoubtedly some of the most successful public health interventions of all time, preventing approximately 2.5 million deaths annually [1]. However, vaccine-preventable diseases evolve and emerge, and novel vaccines and vaccination strategies are developed in response. In order to maintain safe and effective vaccination programs, it is critical that those responsible for making vaccine policy and clinical practice guidelines act promptly to develop and implement evidence-based recommendations. A review of the quality of the evidence relied upon, and identification of the determinants of reduced quality, could help guide researchers, and research funders, on how vaccine studies could be better designed in order to maximise the value of the evidence produced.

In Australia, the Australian Technical Advisory Group on Immunisation (ATAGI) develops evidence-based vaccine clinical practice recommendations that are published in the Australian Immunisation Handbook [2]. ATAGI develops recommendations according to guidelines developed by the National Health and Medical Research Council (NHMRC) [3] and documents their decision-making processes transparently [4]. At the core of the NHMRC guidelines lies the Grading of Recommendations Assessment, Development and Evaluation (GRADE) methodology, a transparent, systematic and methodical framework for developing and presenting summaries of evidence and its certainty [5–8]. When a new clinical practice question arises, for example with the licensure of a new vaccine, ATAGI undertakes a GRADE assessment to review and assess the relevant evidence with technical support provided by the National Centre for Immunisation Research and Surveillance (NCIRS) [4]. The GRADE assessments are made publicly accessible on the NCIRS website [9–16].

Similarly, the Advisory Committee on Immunization Practices (ACIP) and the Joint Committee on Vaccination and Immunisation (JCVI) develop vaccine clinical practice recommendations for the United States of America and the United Kingdom, respectively. ACIP also implements the GRADE methodology as described in the U.S. ACIP Handbook for Developing Evidence-based Recommendations [18], documents their decision-making processes [19] and publicly reports their GRADE assessments and corresponding recommendations on their website and the Morbidity and Mortality Weekly Report [19]. Similarly, since 2008, the World Health Organization (WHO) has used the GRADE approach for the development of their recommendations on immunisation policies [20]. JCVI publish their recommendations in the Immunisation against Infectious Disease handbook (the "Green Book") [17] but the process for developing these recommendations is undocumented.

The GRADE working group has developed and refined the GRADE methodology since its inception in 2000 and has documented their process in the GRADE Handbook [21]. The GRADE method allows decision-makers (e.g., policy-makers and guideline developers) to assess the current state of relevant clinical research evidence to inform their decision-making transparently (i.e., guideline or recommendation development). First, the assessors must clearly define the research question. This is typically framed using the PICO (population, intervention, comparator and outcomes) format, a practice well-established in the evidence-based medicine community [22–23]. Next, a systematic review is conducted to identify all relevant clinical studies that may provide evidence to answer the research question. For each outcome specified in the PICO question, evidence is collated across the identified sources and a certainty of evidence grade is systematically determined with the following possible classifications: *very low*, *low*, *moderate* or *high* [21]. In general, the default classification for evidence certainty obtained from randomised clinical trials (RCTs) is *high*, but this may be downgraded for reasons such as *risk of bias*, *inconsistency*, *indirectness* or *imprecision*, or upgraded due to a *large effect magnitude*, *clear dose-response gradient* or *reverse residual confounding* [21]. The default classification for evidence certainty obtained from observational studies is *low,* and this may be downgraded or upgraded for the same reasons. After each outcome has been graded, an overall certainty of evidence grade is assigned for the research question and the assessors proceed with either a *strong* or *weak* recommendation *for* or *against* the proposed intervention, which is informed by the overall grade.

Here we describe and summarise the publicly available GRADE assessments and review the quality of research evidence that has been relied upon to inform vaccine clinical practice recommendations in Australia. We do not comment on the decision-making process followed by ATAGI, nor the recommendations made or the individual GRADE assessments themselves. Instead, we focus on the quality of the evidence used to inform vaccine recommendations in order to determine whether there is a prima facie need to improve evidence quality and, if so, to provide insights into how evidence quality could be improved. We do not concern ourselves with the magnitude or direction of the evidence, nor the subsequent recommendations made by ATAGI.

## Material and methods

We systematically reviewed ATAGI’s publicly accessible GRADE assessments for the use of vaccines across the following disease areas: cholera, diphtheria, tetanus and pertussis (DTP), human papillomavirus (HPV), influenza, meningococcal, pneumococcal, rabies and varicella zoster virus (zoster) [9–16]. All GRADE assessments available on the NCIRS website in July 2024 were included (note that the GRADE methodology was adopted by ATAGI for new or updated recommendations in July 2020; recommendations made prior to this have not been considered). In some instances, multiple GRADE assessments were conducted within a disease area, each answering a different research question.

The raw data was collated, stored and analysed in R version 4.4.0 [24] and is available, in addition to the analysis code, at https://github.com/michaeldymock25/quality-of-evidence. For each GRADE assessment, we recorded the certainty of evidence classification for each outcome and overall, in addition to the reasons for downgrade or upgrade (where applicable).

Outcomes containing the search string “adverse events|fever|pain|fatigue|cardiovascular” were categorised as safety outcomes, otherwise they were categorised as efficacy outcomes. Research questions containing the search strings “infants”, “children aged 2-17”, “children and adults”, “adolescents”, “over 18”, “over 50|over 65|over 70” were categorised into age groups *infants*, *children and adolescents*, *children and adults*, *adolescents*, *adults and older adults*, respectively. Similarly, research questions containing the search strings “standard risk|without underlying risk|immunocompetent” and “increased risk|with underlying risk|immunocompromised|high risk of exposure|indicated to receive rabies pre-exposure” were categorised into population risk groups *standard risk of disease* and *increased risk of disease*, respectively. Finally, research questions containing the search strings “non-indigenous” and “indigenous” were categorised into Indigenous status groups *non-Indigenous* and *Indigenous*, respectively. Research questions that were indifferent to age, population risk or ethnicity were excluded from the age group, population risk group and Indigenous status group analyses, respectively. Results are summarised as counts and percentages and visualised as bar and line charts.

## Results

GRADE assessments for 25 research questions across 8 diseases were reviewed. Details on the individual PICO questions and the outcomes assessed are provided in Table 1 and Table 2, respectively. The GRADE assessments within the cholera, DTP, HPV, meningococcal, pneumococcal and rabies disease areas were each initiated following (or in anticipation of) approvals or recommendations of new vaccines and/or schedules by the Therapeutic Goods Administration, WHO, ACIP or the Pharmaceutical Benefits Advisory Committee within the respective disease areas. The reasons for initiating GRADE assessments for influenza and zoster vaccines were not stated. The GRADE assessments considered the most up to date clinical research evidence where most of the studies included were published within the last decade.

**Table 1.**
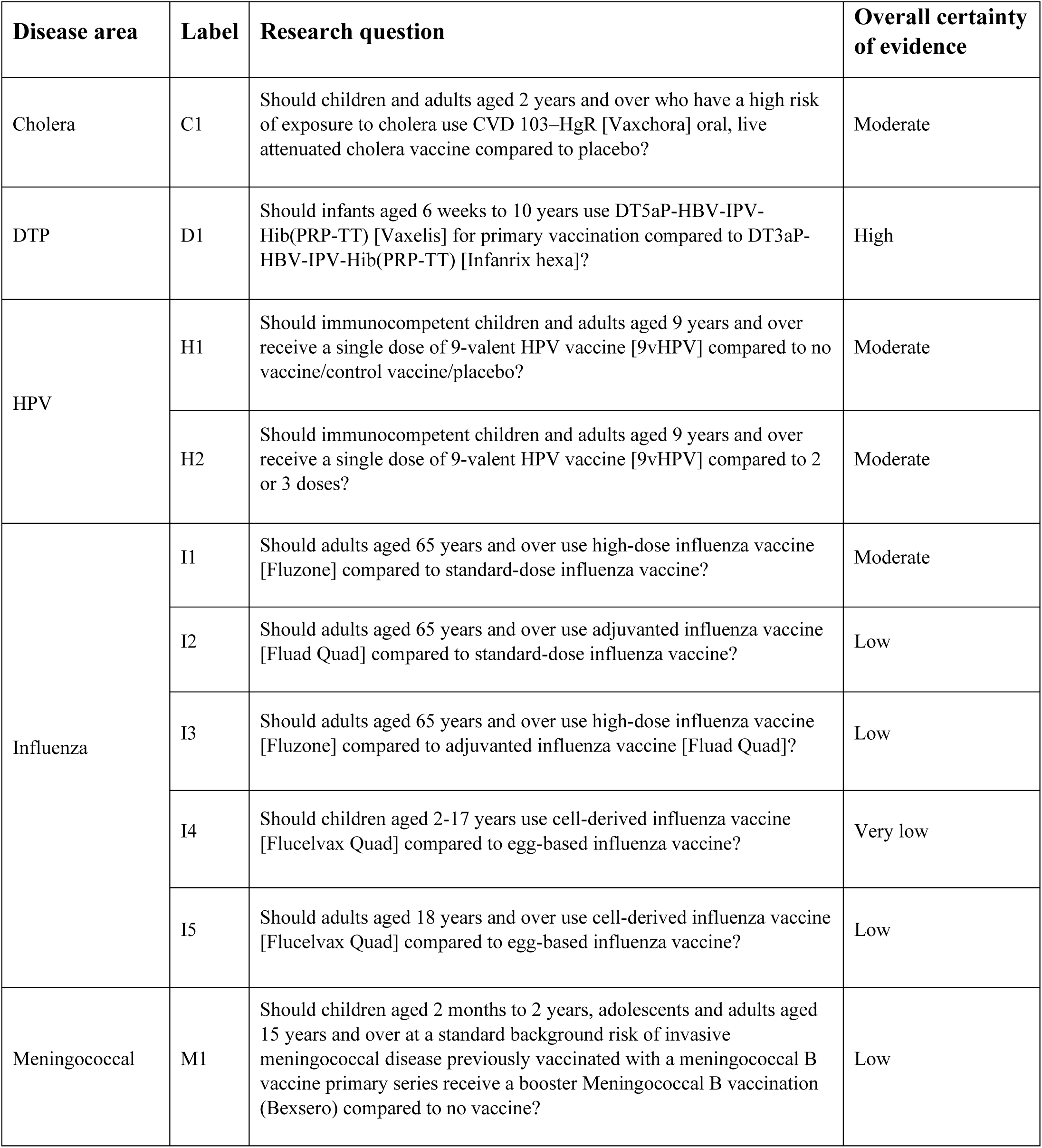
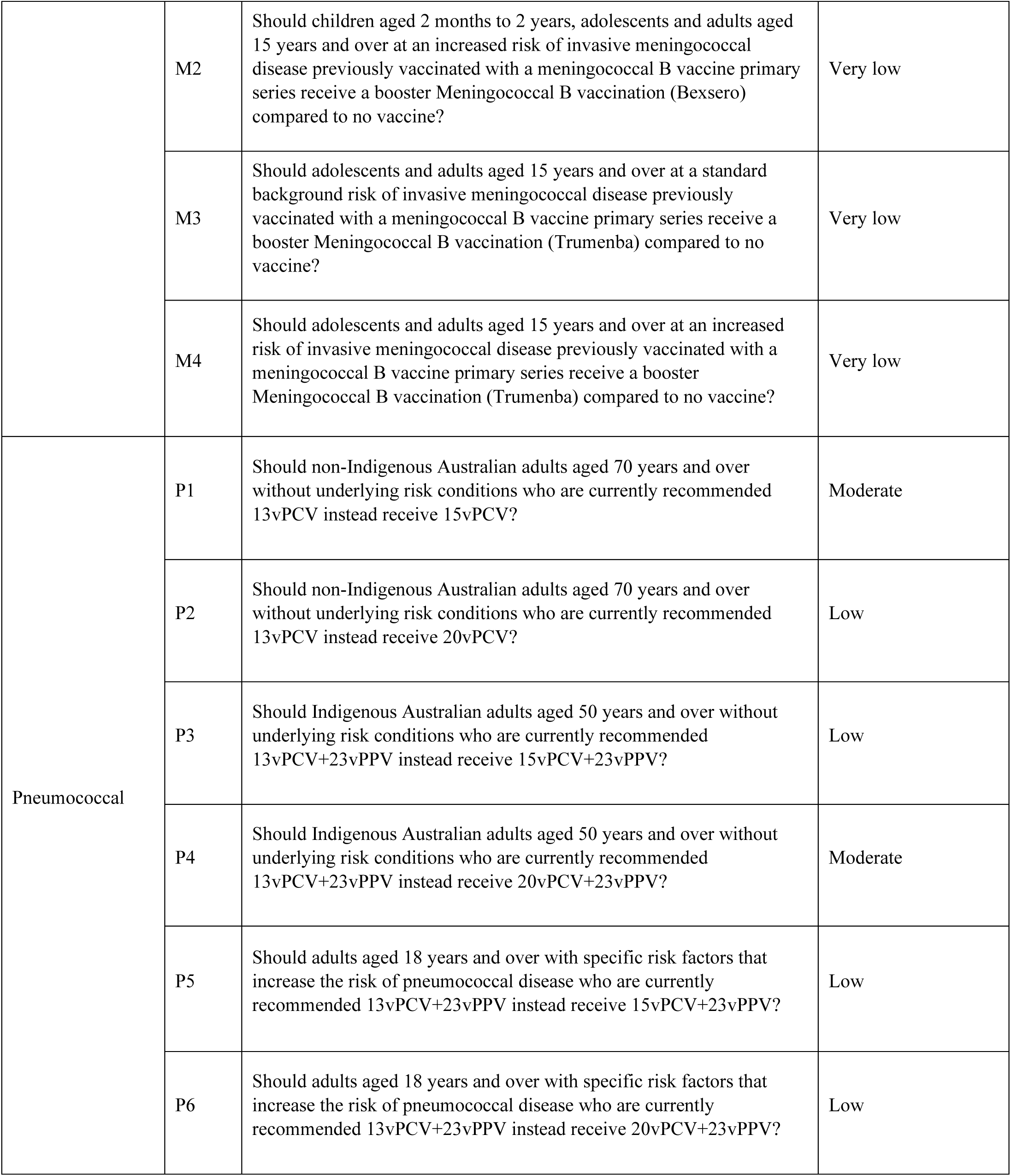
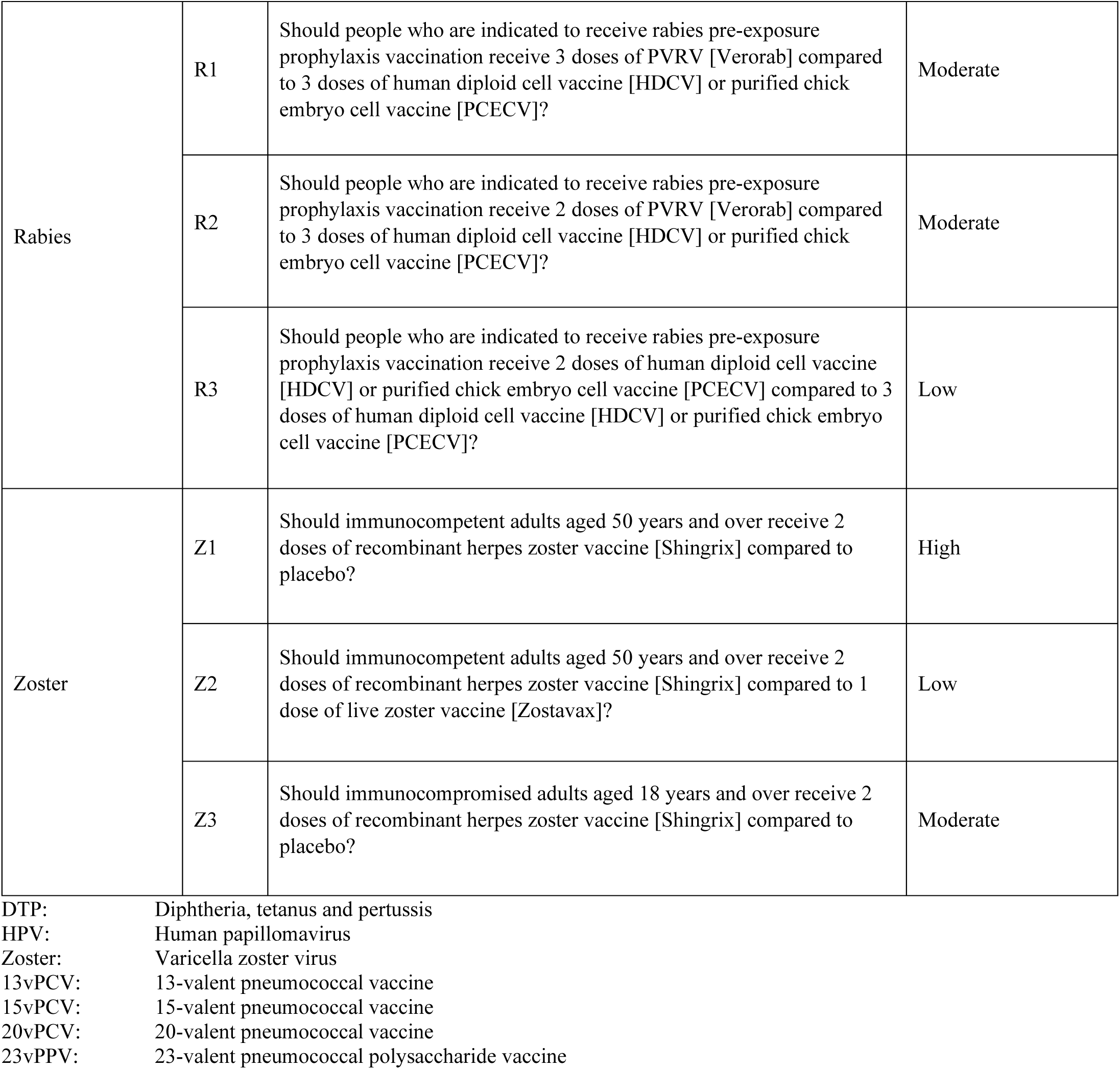
Research questions considered for each disease area. Each research question corresponds to a separate GRADE assessment. Questions have been phrased to make the populations, interventions and comparators clear.

| Disease area | Label | Research question | Overall certainty of evidence |
| --- | --- | --- | --- |
| Cholera | C1 | Should children and adults aged 2 years and over who have a high risk of exposure to cholera use CVD 103–HgR [Vaxchora] oral, live attenuated cholera vaccine compared to placebo? | Moderate |
| DTP | D1 | Should infants aged 6 weeks to 10 years use DT5aP-HBV-IPV-Hib(PRP-TT) [Vaxelis] for primary vaccination compared to DT3aP-HBV-IPV-Hib(PRP-TT) [Infanrix hexa]? | High |
| HPV | H1 | Should immunocompetent children and adults aged 9 years and over receive a single dose of 9-valent HPV vaccine [9vHPV] compared to no vaccine/control vaccine/placebo? | Moderate |
|  | H2 | Should immunocompetent children and adults aged 9 years and over receive a single dose of 9-valent HPV vaccine [9vHPV] compared to 2 or 3 doses? | Moderate |
| Influenza | I1 | Should adults aged 65 years and over use high-dose influenza vaccine [Fluzone] compared to standard-dose influenza vaccine? | Moderate |
|  | I2 | Should adults aged 65 years and over use adjuvanted influenza vaccine [Fluad Quad] compared to standard-dose influenza vaccine? | Low |
|  | I3 | Should adults aged 65 years and over use high-dose influenza vaccine [Fluzone] compared to adjuvanted influenza vaccine [Fluad Quad]? | Low |
|  | I4 | Should children aged 2-17 years use cell-derived influenza vaccine [Flucelvax Quad] compared to egg-based influenza vaccine? | Very low |
|  | I5 | Should adults aged 18 years and over use cell-derived influenza vaccine [Flucelvax Quad] compared to egg-based influenza vaccine? | Low |
| Meningococcal | M1 | Should children aged 2 months to 2 years, adolescents and adults aged 15 years and over at a standard background risk of invasive meningococcal disease previously vaccinated with a meningococcal B vaccine primary series receive a booster Meningococcal B vaccination (Bexsero) compared to no vaccine? | Low |
|  | M2 | Should children aged 2 months to 2 years, adolescents and adults aged 15 years and over at an increased risk of invasive meningococcal disease previously vaccinated with a meningococcal B vaccine primary series receive a booster Meningococcal B vaccination (Bexsero) compared to no vaccine? | Very low |
|  | M3 | Should adolescents and adults aged 15 years and over at a standard background risk of invasive meningococcal disease previously vaccinated with a meningococcal B vaccine primary series receive a booster Meningococcal B vaccination (Trumenba) compared to no vaccine? | Very low |
|  | M4 | Should adolescents and adults aged 15 years and over at an increased risk of invasive meningococcal disease previously vaccinated with a meningococcal B vaccine primary series receive a booster Meningococcal B vaccination (Trumenba) compared to no vaccine? | Very low |
| Pneumococcal | P1 | Should non-Indigenous Australian adults aged 70 years and over without underlying risk conditions who are currently recommended 13vPCV instead receive 15vPCV? | Moderate |
|  | P2 | Should non-Indigenous Australian adults aged 70 years and over without underlying risk conditions who are currently recommended 13vPCV instead receive 20vPCV? | Low |
|  | P3 | Should Indigenous Australian adults aged 50 years and over without underlying risk conditions who are currently recommended 13vPCV+23vPPV instead receive 15vPCV+23vPPV? | Low |
|  | P4 | Should Indigenous Australian adults aged 50 years and over without underlying risk conditions who are currently recommended 13vPCV+23vPPV instead receive 20vPCV+23vPPV? | Moderate |
|  | P5 | Should adults aged 18 years and over with specific risk factors that increase the risk of pneumococcal disease who are currently recommended 13vPCV+23vPPV instead receive 15vPCV+23vPPV? | Low |
|  | P6 | Should adults aged 18 years and over with specific risk factors that increase the risk of pneumococcal disease who are currently recommended 13vPCV+23vPPV instead receive 20vPCV+23vPPV? | Low |
| Rabies | R1 | Should people who are indicated to receive rabies pre-exposure prophylaxis vaccination receive 3 doses of PVRV [Verorab] compared to 3 doses of human diploid cell vaccine [HDCV] or purified chick embryo cell vaccine [PCECV]? | Moderate |
|  | R2 | Should people who are indicated to receive rabies pre-exposure prophylaxis vaccination receive 2 doses of PVRV [Verorab] compared to 3 doses of human diploid cell vaccine [HDCV] or purified chick embryo cell vaccine [PCECV]? | Moderate |
|  | R3 | Should people who are indicated to receive rabies pre-exposure prophylaxis vaccination receive 2 doses of human diploid cell vaccine [HDCV] or purified chick embryo cell vaccine [PCECV] compared to 3 doses of human diploid cell vaccine [HDCV] or purified chick embryo cell vaccine [PCECV]? | Low |
| Zoster | Z1 | Should immunocompetent adults aged 50 years and over receive 2 doses of recombinant herpes zoster vaccine [Shingrix] compared to placebo? | High |
|  | Z2 | Should immunocompetent adults aged 50 years and over receive 2 doses of recombinant herpes zoster vaccine [Shingrix] compared to 1 dose of live zoster vaccine [Zostavax]? | Low |
|  | Z3 | Should immunocompromised adults aged 18 years and over receive 2 doses of recombinant herpes zoster vaccine [Shingrix] compared to placebo? | Moderate |

DTP: Diphtheria, tetanus and pertussis HPV: Human papillomavirus Zoster: Varicella zoster virus 13vPCV: 13-valent pneumococcal vaccine 15vPCV: 15-valent pneumococcal vaccine 20vPCV: 20-valent pneumococcal vaccine 23vPPV: 23-valent pneumococcal polysaccharide vaccine

**Table 2.** Summary of outcomes used in the GRADE assessments by disease area. Outcome summaries apply to all research questions (GRADE assessments) within a disease area noting that individual assessments may have considered more specific definitions (e.g., included a time window for outcome ascertainment).

| **Disease Area** | **Efficacy Outcomes** | **Safety Outcomes** |
| --- | --- | --- |
| Cholera | Mild, moderate or severe cholera diarrhoea<br>Serum vibriocidal antibody seroconversion | Serious adverse events<br>Systemic adverse events |
| DTP | Antibody titres | Serious adverse events<br>Systemic adverse events<br>Local adverse events<br>Adverse events of special interest<br>Fever events |
| HPV | HPV infection<br>Genital infection<br>Cervical intraepithelial neoplasia grade 3 infection<br>Antibody titres | Serious adverse events<br>Systemic adverse events<br>Local adverse events |
| Influenza | All-cause mortality<br>Influenza- or pneumonia-associated mortality<br>All-cause hospitalisation<br>Influenza-, pneumonia- or respiratory-related hospitalisation<br>Hospitalisation for pneumonia, stroke or myocardial infarction<br>Influenza- or pneumonia-related office visits<br>Laboratory-confirmed influenza<br>Reverse transcription PCR- or culture-confirmed influenza<br>PCR-confirmed influenza A<br>PCR-confirmed influenza B<br>Influenza-like illness | Serious adverse events<br>Systemic adverse events<br>Local adverse events<br>Adverse events of special interest<br>Cardiovascular events |
| Meningococcal | Antibody titres | Systemic adverse events<br>Local adverse events<br>Unsolicited adverse events<br>Fever events<br>Pain events |
|  |  | Fatigue events |
| Pneumococcal | Antibody titres | Serious adverse events<br>Systemic adverse events<br>Local adverse events |
| Rabies | Rabies virus neutralising antibody seroconversion | Vaccine-related serious adverse events<br>Systemic adverse events<br>Local adverse events |
| Zoster | Herpes zoster infection<br>Post-herpetic neuralgia<br>Herpes zoster-related hospitalisation<br>Immune-mediated disease<br>Humoral immunogenicity<br>Cell-mediated immunogenicity | Serious adverse events<br>Systemic adverse events<br>Local adverse events<br>Unsolicited adverse events |

DTP: Diphtheria, tetanus and pertussis HPV: Human papillomavirus PCR: Polymerase chain reaction Zoster: Varicella zoster virus

Across the 25 research questions, 189 separate outcomes were assessed using the eligible studies; of these outcomes, 43 (22.8%), 38 (20.1%), 68 (36.0%) and 40 (21.2%) were classified as being informed by *very low*, *low*, *moderate* and *high* certainty of evidence, respectively. Figure 1 displays the breakdown of certainty of evidence classifications for the outcomes across each research question within each disease area. The certainty of evidence varied across disease areas. Cholera, DTP and HPV assessments were informed by *moderate* or *high* certainty evidence only.

**Figure 1.**
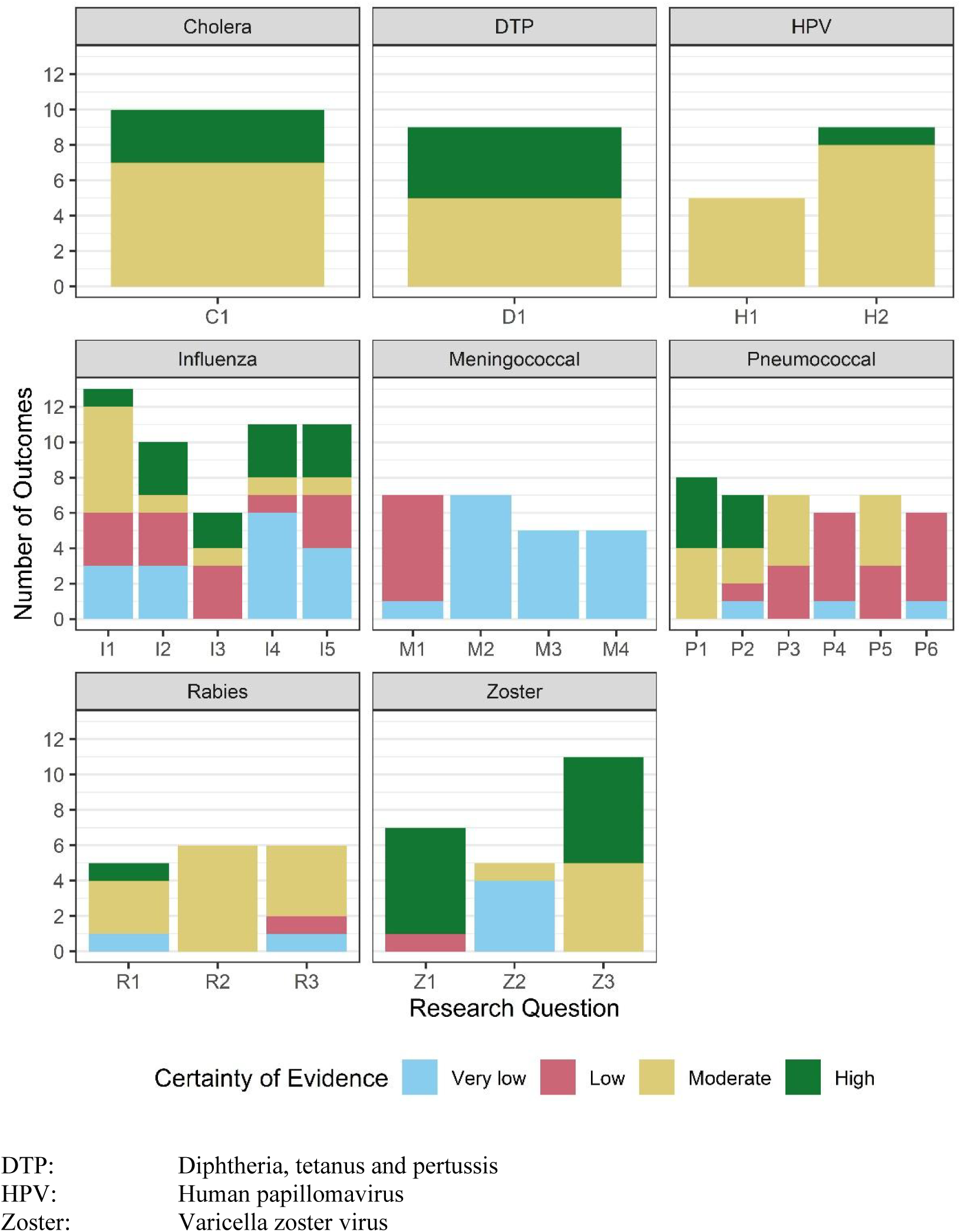
GRADE certainty of evidence classifications for individual outcomes by research question within disease area. Research question labels correspond to the labels found in Table 1.

Meningococcal assessments were informed by *very low* or *low* certainty evidence. Influenza, pneumococcal, rabies and zoster assessments were informed by evidence across the range of certainty classifications. Figure 2 shows similar variation between the certainty of evidence classifications across disease areas when the outcomes are categorised into efficacy (109 outcomes) and safety (80 outcomes). Overall, 4 (16%), 10 (40%), 9 (36%) and 2 (8%) research questions across the disease areas had their overall certainty of evidence classified as *very low*, *low*, *moderate* and *high*, respectively. The distribution of overall certainty of evidence classifications varied across disease areas but did not appear to vary according to the research questions’ focus on age, population risk or Indigenous status (Table 1).

**Figure 2.**
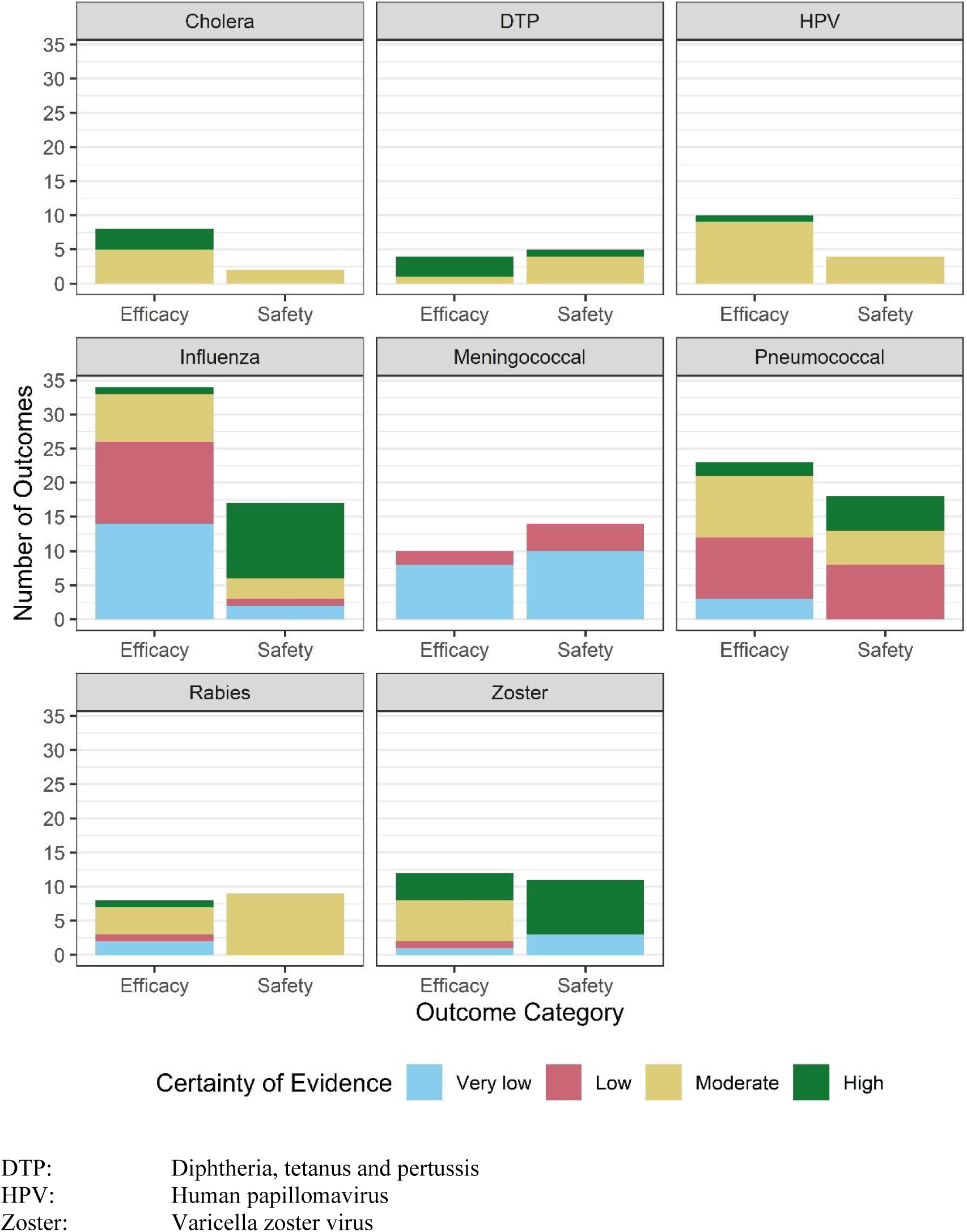
GRADE certainty of evidence classifications for individual outcomes by outcome category (efficacy or safety) within disease area.

Of the 79 (41.8%) outcomes that had their certainty of evidence downgraded due to risk of bias, the most common reasons were potential confounding (28; 35.4%), single arm studies (24; 30.4%) and selective outcome reporting (7; 8.9%). There were 9 (4.8%) outcomes with downgraded certainty of evidence for inconsistency due to variability in the results between studies (6; 66.7%) or wide prediction intervals (3; 33.3%); inconsistency was not assessed for 48 (25.4%) outcomes due to only one study being included in the comparison. Of the 70 (37.0%) outcomes with downgraded certainty of evidence for indirectness, most reasons were related to differences between the populations included in the studies and the population defined in the research question (38; 54.3%) and/or differences between other study elements (e.g. intervention or comparator schedule) and those defined in the research question (34; 48.6%), although there could be multiple reasons for downgrading. Finally, there were 81 (42.9%) outcomes with downgraded certainty of evidence for imprecision due to small sample sizes (29; 35.8%), wide confidence intervals or “statistically non-significant” results (23; 28.4%), a low number of events (22; 27.2%) or the studies being underpowered (7; 8.6%). Figure 3 displays the number of levels each outcome was downgraded for each assessment category (risk of bias, inconsistency, indirectness or imprecision) for each disease area. Figures S1-S6 in the Supplementary Materials show how each outcome was downgraded for each assessment category and each research question for each disease area (excluding cholera and DTP as each of these disease areas only have one research question).

**Figure 3.**
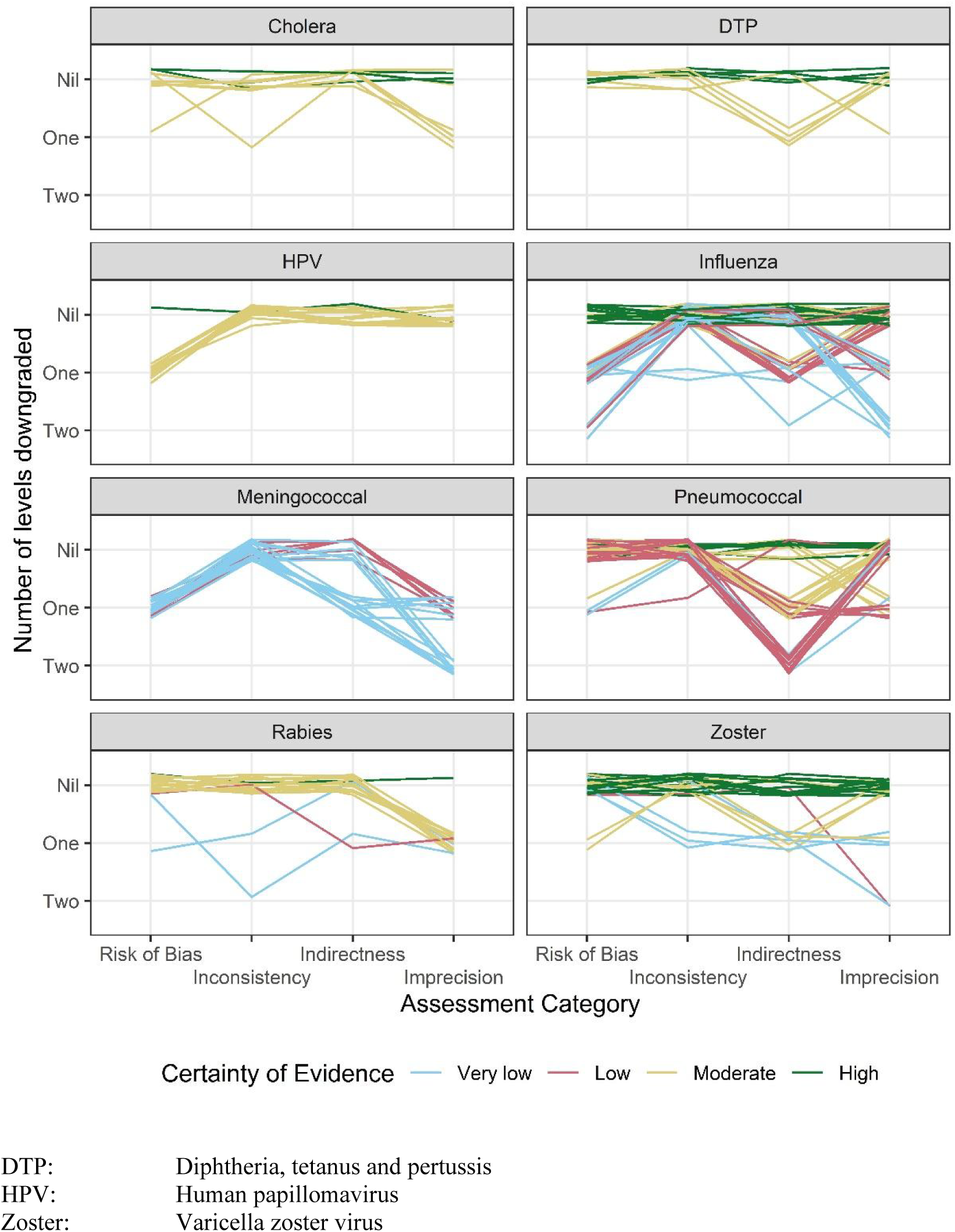
Certainty of evidence classification for each outcome across each assessment category within each disease area. Outcomes classified High, Moderate, Low and Very Low have been downgraded zero, one, two and three or more levels, respectively.

There were 6 outcomes where the certainty of evidence classification was upgraded due to a large effect magnitude. However, for one of the outcomes, risk of bias, inconsistency, indirectness and imprecision were all graded serious and so the certainty of evidence was classified *very low* even after accounting for the large effect magnitude. The large effect magnitude for the other 5 outcomes similarly had no impact as the certainty of evidence was already classified as *high*.

## Discussion

In this review, we found that around half (56%) of the GRADE assessments informing Australian vaccine practice guidelines received an overall certainty of evidence classification of *low* or *very low*, which likely impacts upon ATAGI’s ability to develop evidence-based recommendations. Reasons for downgrading the certainty of evidence for individual outcomes included potential for confounding, mismatch between the question-specific versus the study-specific population and design (e.g., interventions and schedules), and uncertainty in the effect estimation (e.g., wide confidence intervals, a low number of events or concerns regarding statistical significance).

While it is always preferable to base policy and practice recommendations on higher rather than lower certainty of evidence, we note that it is neither necessary nor desirable for clinical practice to always be guided by the highest certainty of evidence. The cost to generate evidence via research must be proportionate to the value of the information gained. Where the risks of vaccination are small, having high precision on the size of the protective effect may be unnecessary. Also, for some vaccine-preventable conditions like meningococcal disease, the rarity of infection makes it infeasible to undertake definitive trials that directly measure the effect of vaccines against infection, so it may be necessary to instead base recommendations on immune responses or other indirect evidence. Nonetheless, our review indicates that there are still likely to be opportunities to improve the certainty of evidence informing vaccine recommendations, for example by designing studies that are more inclusive of subpopulations, that are inclusive of vaccine schedules under programmatic consideration (including dose-sparing schedules), and that consider the cost-effectiveness of vaccine strategies and provide granular probabilistic outputs to assist decision-making. The use of contemporary methods to minimise confounding and selection bias in the analysis of observational data, in conjunction with improved standards of reporting, should also be considered by researchers to improve the quality of evidence available for GRADE assessments.

GRADE assessments are now used routinely to guide vaccine clinical practice recommendations in Australia and USA. GRADE has also been used to assess the certainty of evidence for COVID-19 vaccine uptake by ethnic minority individuals in the United Kingdom [25], risk management of allergic reactions to COVID-19 vaccines [26–27], use of the two-dose varicella vaccine in China [28], strategies for addressing vaccine hesitancy [29], and vaccination against rotavirus [30] and hepatitis B [31] in Germany. While the intended purpose of GRADE is to guide the translation of evidence into recommendations, in this review we have utilised it instead to identify potential limitations of the evidence used to inform vaccine recommendations, and to identify opportunities for researchers to improve the quality of evidence relied upon. With increased adoption of the GRADE methodology, there are efforts to expand and improve the GRADE approach. Guidance developed by the GRADE Public Health Group [32] addresses the remaining limitations in using GRADE to develop vaccine clinical practice recommendations, such as the prioritisation of outcomes and inclusion of perspectives from diverse stakeholders.

One challenge that assessors face, especially when asking questions where evidence is scarce (e.g., for rare populations), is a lack of any high-quality evidence. Although GRADE does allow for the consideration of lower quality observational research evidence in situations where RCTs are unfeasible or unavailable, such as post-marketing vaccine safety surveillance evidence generated by AusVaxSafety [33], it may be impossible to (fully) alleviate bias in the available evidence, let alone quantify its magnitude.

In this manuscript we presented descriptive summaries of the GRADE assessments that have been used by ATAGI to develop new and updated vaccine clinical practice recommendations in Australia from July 2020 to July 2024. Due to this time period restriction, bias in the selection of assessments considered in this work cannot be evaluated and so we avoid making broad generalised interpretations of the results. We do not attempt to interpret the comparisons of evidence certainty across disease areas and populations as these may be disproportionately affected by external factors such as the feasibility of research and acknowledge this as a limitation. Furthermore, we acknowledge that the certainty of evidence classifications, although crucial, are not the only component of the evidence-to-decision framework used by ATAGI when making vaccine clinical practice recommendations. The priority of the research question, acceptability to stakeholders and feasibility are other important factors considered [3].

Clinical research should aim to impact and inform policy and practice, either directly by addressing policy-specific questions, or indirectly by contributing to an evolving evidence base. It stands to reason that research (in particular research that is publicly funded) should be designed to answer the research questions of policy-makers most efficiently. In the context of vaccine clinical practice recommendations, researchers must consider how to improve the quality of evidence used by policy-makers. Acknowledging that clinical research is subject to external constraints from limitations in funding and recruitment, it will be difficult to completely remedy all issues related to potential for bias and uncertainty reduction in estimation. One way to improve the quality of evidence which may have few or no resource implications, would be to better align future studies with policy questions. However, this would require policy-makers to be involved in setting research questions *before* the evidence is generated, rather than afterwards.

## Author contributions

MD drafted the primary version of this manuscript, conducted the data curation and investigation and developed the software and visualisation. JM, KM and TS assisted with the study conceptualisation, reviewed and edited the manuscript and provided supervision for MD. All authors attest they reviewed and approved the final version for publication and met the ICMJE criteria for authorship.

## Data Availability

All data used in this work are available online at https://github.com/michaeldymock25/quality-of-evidence.

## Acknowledgements

The authors express their gratitude to Clayton Chiu and Zhicheng (Jeff) Wang for their extensive comments on the draft manuscript. MD is supported by a NHMRC Postgraduate Research Award (GNT2022557), an AusTriM Postgraduate Top-up Scholarship, a Stan and Jean Perron Top-up Scholarship and a Statistical Society of Australia PhD Top-up Scholarship. TS is supported by a MRFF Investigator Grant (MRF1195153).

## References

[1]. World Health Organization, UNICEF and World Bank. State of the world’s vaccines and immunization (3rd edition). *Geneva: World Health Organization*. 2009. https://iris.who.int/handle/10665/44169 [accessed 7 May 2024].

[2]. Australian Technical Advisory Committee on Immunisation. Australian Immunisation Handbook. *Australian Government Department of Health and Aged Care*. 2022. https://immunisationhandbook.health.gov.au [accessed 7 May 2024].

[3] National Health and Medical Research Council. Guidelines for Guidelines Handbook. National Health and Medical Research Council. 2016. https://www.nhmrc.gov.au/guidelinesforguidelines [accessed 7 May 2024].

[4]. Australian Technical Advisory Committee on Immunisation. ATAGI decision-making process for developing clinical recommendations in the Australian Immunisation Handbook. *Australian Government Department of Health and Aged Care*. 2022. https://immunisationhandbook.health.gov.au/resources/publications/atagi-decision-making-process-for-developing-clinical-recommendations-in-the-australian-immunisation-handbook [accessed 7 May 2024].

[5] Guyatt GH, Oxman AD, Vist GE, Kunz R, Falck-Ytter Y, Alonso-Coello P, Schünemann HJ. GRADE: an emerging consensus on rating quality of evidence and strength of recommendations. BMJ. 2008; 336 (7650): 924–6. 10.1136/bmj.39489.470347.AD.

[6] Guyatt GH, Oxman AD, Kunz R, Vist GE, Falck-Ytter Y, Schünemann HJ. What is quality of evidence and why is it important to clinicians? BMJ. 2008; 336 (7651): 995–8. 10.1136/bmj.39490.551019.BE.

[7] Guyatt GH, Oxman AD, Kunz R, Falck-Ytter Y, Vist GE, Liberati A, Schünemann HJ. Going from evidence to recommendations. BMJ. 2008; 336 (7652): 1049–51. 10.1136/bmj.39493.646875.AE.

[8] Guyatt GH, Oxman AD, Kunz R, Jaeschke R, Helfand M, Liberati A, Vist GE, Schünemann HJ. Incorporating considerations of resources use into grading recommendations. BMJ. 2008; 336 (7654): 1170–3. 10.1136/bmj.39504.506319.80.

[9]. National Centre for Immunisation Research and Surveillance. Cholera vaccines GRADE assessments. *National Centre for Immunisation Research and Surveillance website*. www.ncirs.org.au/cholera-vaccines-grade-assessments-0 [accessed 7 May 2024].

10. National Centre for Immunisation Research and Surveillance. Diphtheria and tetanus toxoids and acellular pertussis (DTPa) containing vaccines GRADE assessments. *National Centre for Immunisation Research and Surveillance website*. www.ncirs.org.au/diphtheria-and-tetanus-toxoids-and-acellular-pertussis-dtpa-containing-vaccines-grade-assessments [accessed 7 May 2024].

11. National Centre for Immunisation Research and Surveillance. Human papillomavirus (HPV) vaccines GRADE assessments. *National Centre for Immunisation Research and Surveillance website*. www.ncirs.org.au/our-work/australian-immunisation-handbook/hpv-grade-assessments [accessed 7 May 2024].

12. National Centre for Immunisation Research and Surveillance. Influenza vaccines GRADE assessments. *National Centre for Immunisation Research and Surveillance website*. www.ncirs.org.au/our-work/australian-immunisation-handbook/influenza-grade-assessments [accessed 7 May 2024].

13. National Centre for Immunisation Research and Surveillance. Meningococcal vaccines GRADE assessments. *National Centre for Immunisation Research and Surveillance website*. www.ncirs.org.au/meningococcal-vaccines-grade-assessments [accessed 7 May 2024].

14. National Centre for Immunisation Research and Surveillance. Pneumococcal vaccines GRADE assessments. *National Centre for Immunisation Research and Surveillance website*. www.ncirs.org.au/pneumococcal-vaccines-grade-assessments [accessed 7 May 2024].

15. National Centre for Immunisation Research and Surveillance. Rabies vaccines GRADE assessments. *National Centre for Immunisation Research and Surveillance website*. www.ncirs.org.au/rabies-vaccines-grade-assessments-0 [accessed 7 May 2024].

[16] National Centre for Immunisation Research and Surveillance. Zoster vaccines GRADE assessments. *National Centre for Immunisation Research and Surveillance website*. www.ncirs.org.au/our-work/australian-immunisation-handbook/zoster-grade-assessments [accessed 7 May 2024].

[17] Ramsay, M. Immunisation against infectious disease. UK Health Security Agency. 2020. www.gov.uk/government/collections/immunisation-against-infectious-disease-the-green-book [accessed 7 May 2024].

[18] Centers for Disease Control and Prevention.. U.S. ACIP Handbook for Developing Evidence-based Recommendations. *Centers for Disease Control and Prevention (CDC),* *Atlanta**, Georgia, United States.* 2024. https://www.cdc.gov/vaccines/acip/recs/grade/about-grade.html [accessed 7 May 2024].

[19] Ahmed F, Temte JL, Campos-Outcalt D, Schünemann HJ. Methods for developing evidence-based recommendations by the advisory committee on immunization practices (ACIP) of the U.S. centers for disease control and prevention (CDC). Vaccine. 2011; 29 (49): 9171–76. 10.1016/j.vaccine.2011.08.005.

[20] Duclos P, Durrheim DN, Reingold AL, Bhutta ZA, Vannice K, Rees H. Developing evidence-based immunization recommendations and GRADE. Vaccine. 2012; 31 (1): 12–19. 10.1016/j.vaccine.2012.02.041.

[21] Schünemann HJ, Brożek J, Guyatt GH, Oxman AD. GRADE Handbook. *Cochrane Training.* 2013. https://training.cochrane.org/resource/grade-handbook [accessed 7 May 2024].

[22] Richardson WS, Wilson MC, Nishikawa J, Hayward RS. The well-built clinical question: a key to evidence-based decisions. American College of Physicians Journal Club. 1995; 123 A12–3. 10.7326/ACPJC-1995-123-3-A12.

[23] Speckman RA, Friedly JL. Asking Structured, Answerable Clinical Questions Using the Population, Intervention/Comparator, Outcome (PICO) Framework. *PM&R The Journal of Injury*, Function and Rehabilitation. 2019; 11 (5): 548–53. 10.1002/pmrj.12116.

[24] R Core Team. R: A language and environment for statistical computing. *R Foundation for Statistical Computing, Vienna*, Austria. 2024. https://www.R-project.org.

[25] Treweek S, Brazzelli M, Crosse A, Daga S, Isaacs T, Sunga R. Using the GRADE evidence to decision framework to reach recommendations together with ethnic minority community organizations: the example of COVID-19 vaccine uptake in the United Kingdom. Journal of Clinical Epidemiology. 2024; 168: 111268. 10.1016/j.jclinepi.2024.111268.

[26] Greenhawt M, Abrams EM, Shaker M, Chu DK, Khan D, Akin C, et al. The Risk of Allergic Reactions to SARS-CoV-2 Vaccines and Recommended Evaluation and Management: A Systematic Review, Meta-Analysis, GRADE Assessment and International Consensus Approach. Journal of Allergy and Clinical Immunology. In Practice. 2021; 9 (10): 3546–67. 10.1016/j.jaip.2021.06.006.

[27] Greenhawt M, Dribin TE, Abrams EM, Shaker M, Chu DK, Golden DBK, et al. Updated guidance regarding the risk of allergic reactions to COVID-19 vaccines and recommended evaluation and management: A GRADE assessment and international consensus approach. Journal of Allergy and Clinical Immunology. 2023; 152 (2): 309–25. 10.1016/j.jaci.2023.05.019.

[28] Zhang Z, Suo L, Pan J, Zhao D, Lu L. Two-dose varicella vaccine effectiveness in China: a meta-analysis and evidence quality assessment. BMC Infectious Diseases. 2021; 21: 543. 10.1186/s12879-021-06217-1.

[29] Jarrett C, Wilson R, O’Leary M, Eckersberger E, Larson HJ. Strategies for addressing vaccine hesitancy - A systematic review. Vaccine. 2015; 33 (34): 4180–90. 10.1016/j.vaccine.2015.04.040.

[30] Koch J, Wiese-Posselt M, Remschmidt C, Wichmann O, Bertelsmann H, Garbe E, et al. Background paper to the recommendation for routine rotavirus vaccination of infants in Germany. Bundesgesundheitsblatt. 2013; 56: 957–84. 10.1007/s00103-013-1777-3.

[31] Harder T, Remschmidt C, Falkenhorst G, Zimmermann R, Hengel H, et al. Background paper to the revised recommendation for hepatitis B vaccination of persons at particular risk and for hepatitis B postexposure prophylaxis in Germany. Bundesgesundheitsblatt. 2013; 56: 1565–76. 10.1007/s00103-013-1845-8.

[32] Boon MH, Thomson H, Shaw B, Akl EA, Lhachimi SK, et al. Challenges in applying the GRADE approach in public health guidelines and systematic reviews: a concept article from the GRADE Public Health Group. Journal of Clinical Epidemiology. 2021; 135: 42–53. 10.1016/j.jclinepi.2021.01.001.

[33] Pillsbury AJ, Glover C, Jacoby P, Quinn HE, Fathima P, Cashman P, et al. Active surveillance of 2017 seasonal influenza vaccine safety: an observational cohort study of individuals aged 6 months and older in Australia. BMJ Open. 2018; **8**: e023263. 10.1136/bmjopen-2018-023263.

