## Supplementary Materials for "What is the quality of evidence informing vaccine clinical practice recommendations in Australia?"

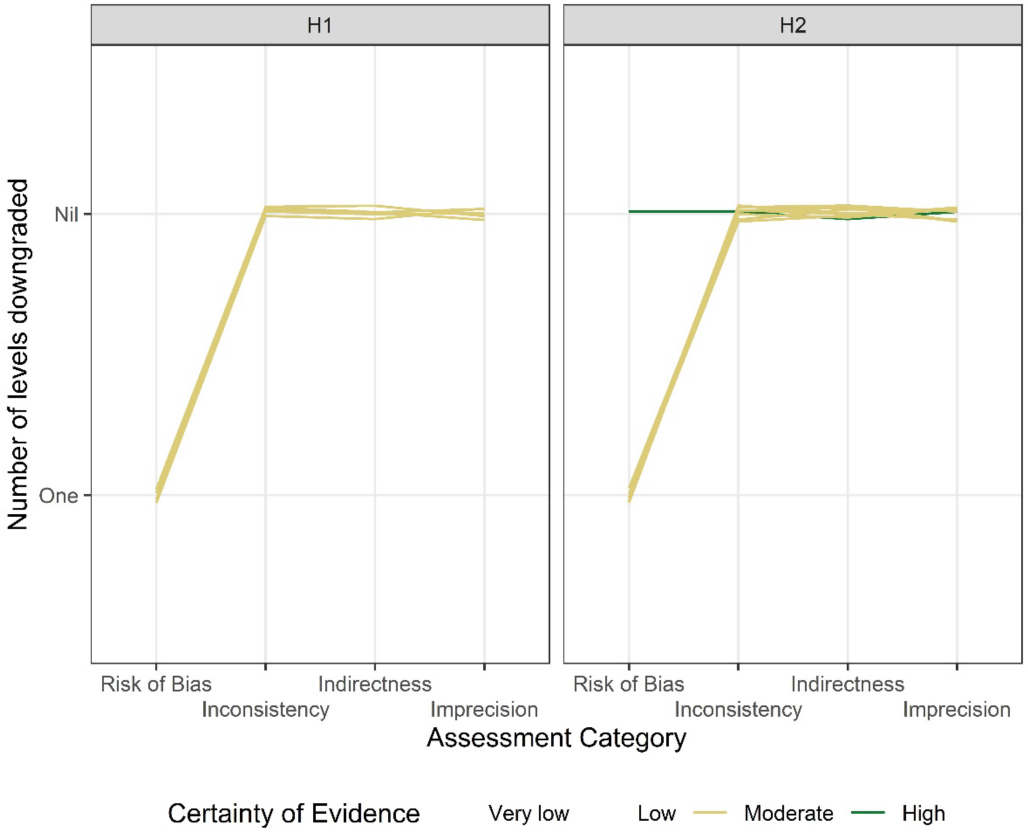


**Figure S1 Certainty of evidence classification for each outcome across each assessment category and research question for human papillomavirus research questions. Outcomes classified High, Moderate, Low and Very Low have been downgraded zero, one, two and three or more levels, respectively.**

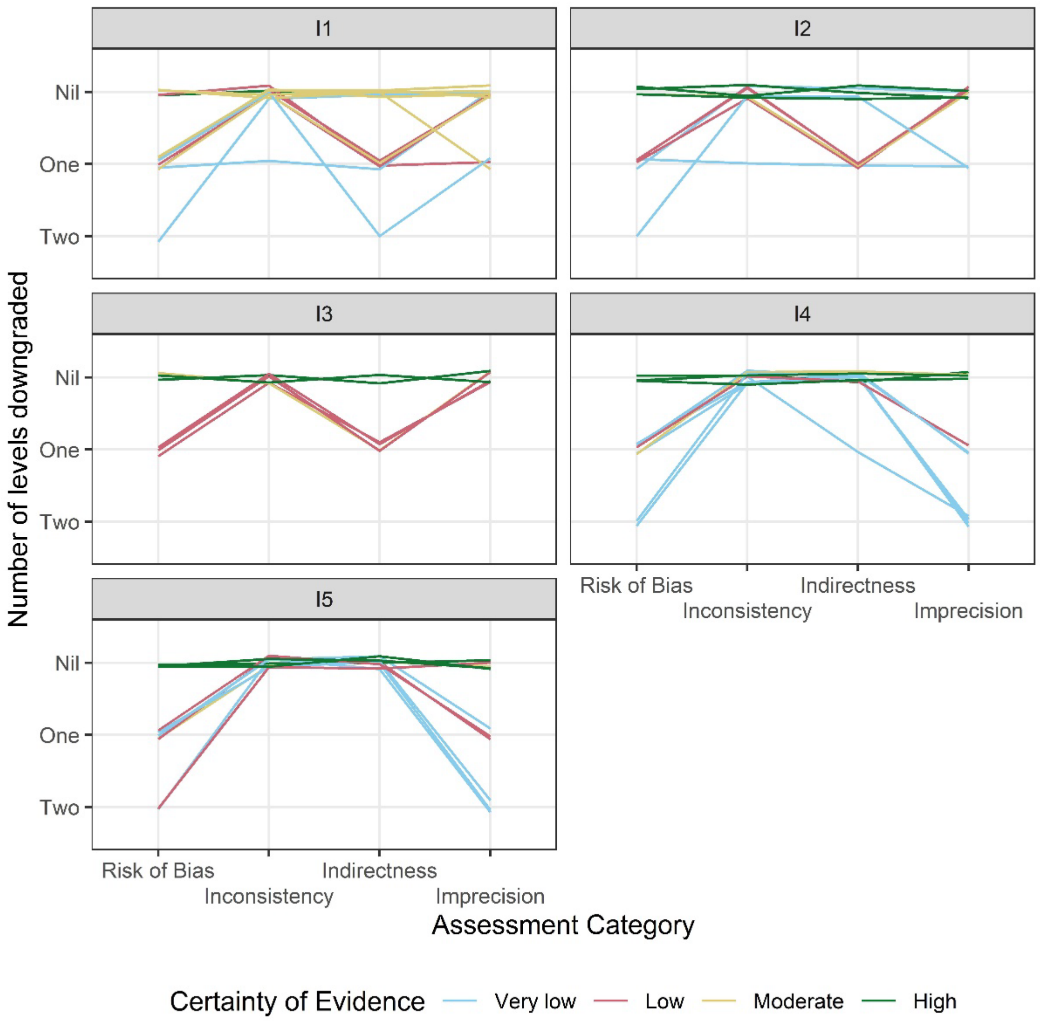


**Figure S2** **Certainty of evidence classification for each outcome across each assessment category and research question for influenza research questions. Outcomes classified High, Moderate, Low and Very Low have been downgraded zero, one, two and three or more levels, respectively.**

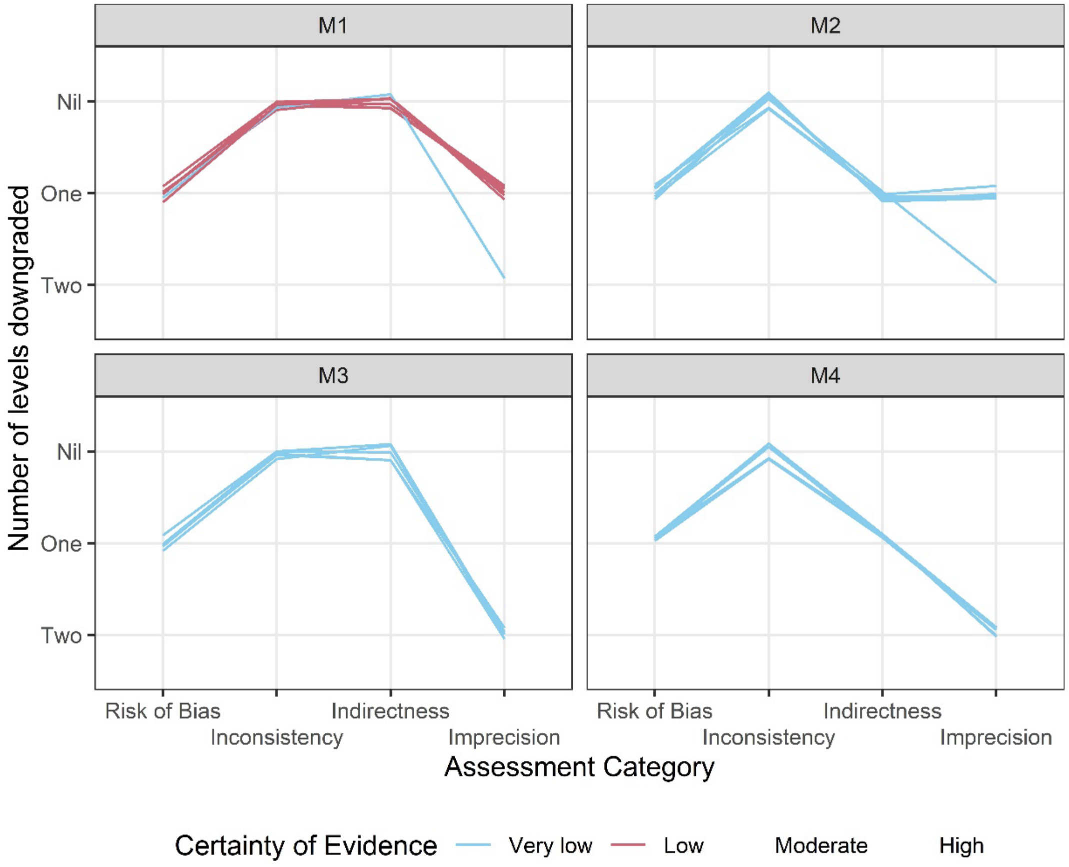


**Figure S3 Certainty of evidence classification for each outcome across each assessment category and research question for meningococcal research questions. Outcomes classified High, Moderate, Low and Very Low have been downgraded zero, one, two and three or more levels, respectively.**

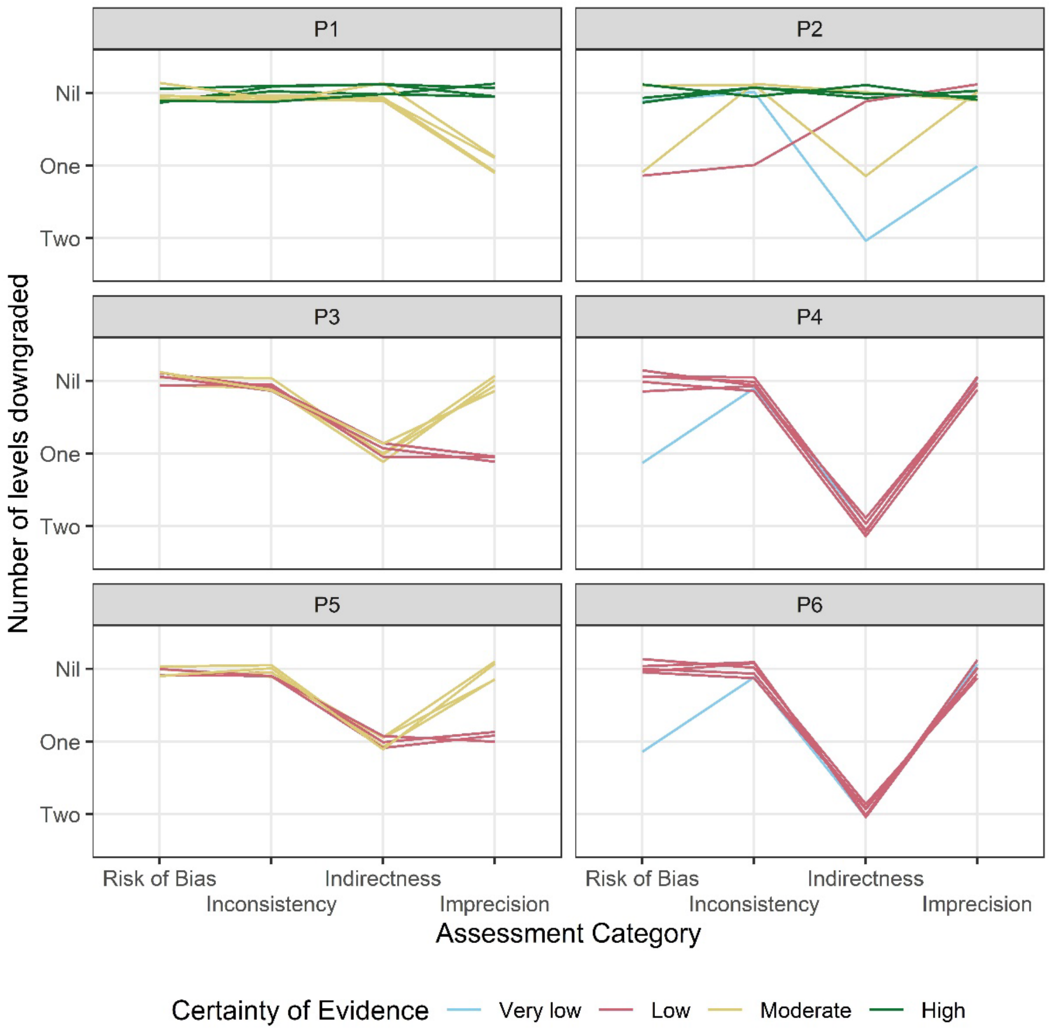


**Figure S4 Certainty of evidence classification for each outcome across each assessment category and research question for pneumococcal research questions. Outcomes classified High, Moderate, Low and Very Low have been downgraded zero, one, two and three or more levels, respectively.**

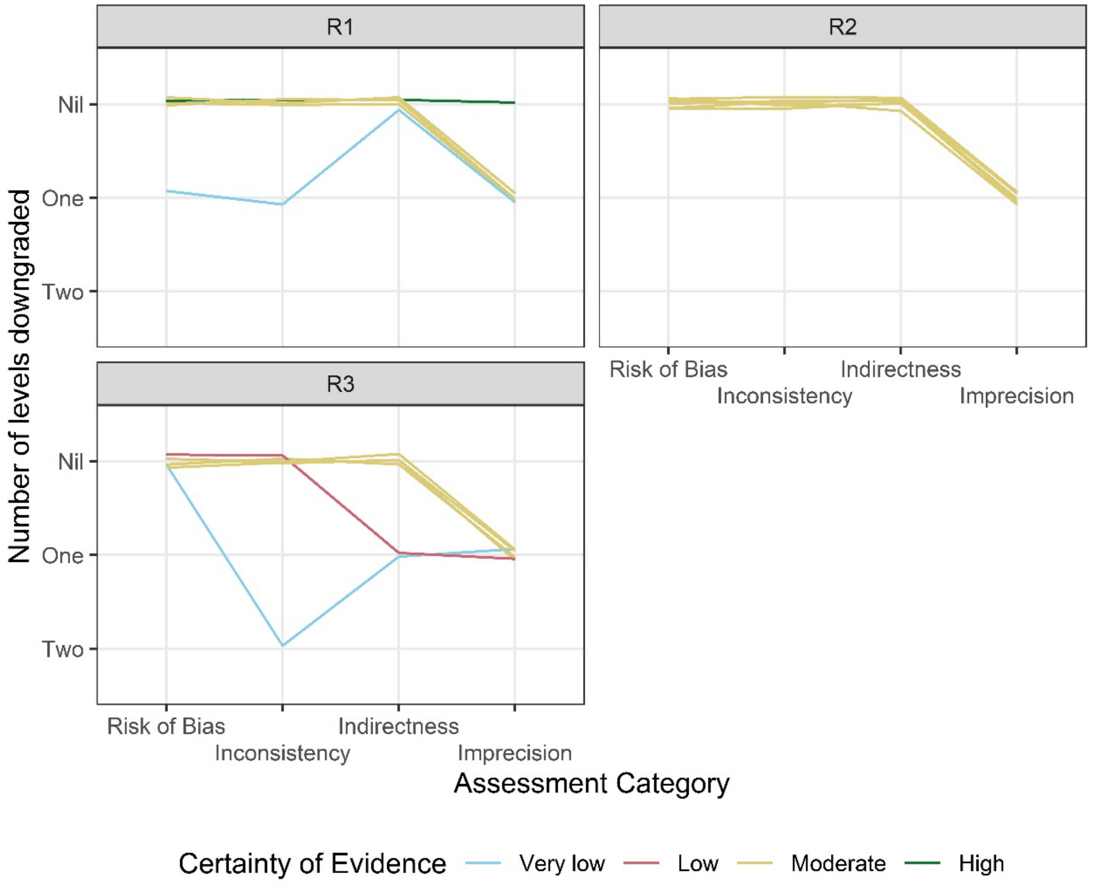


**Figure S5 Certainty of evidence classification for each outcome across each assessment category and research question for rabies research questions. Outcomes classified High, Moderate, Low and Very Low have been downgraded zero, one, two and three or more levels, respectively.**

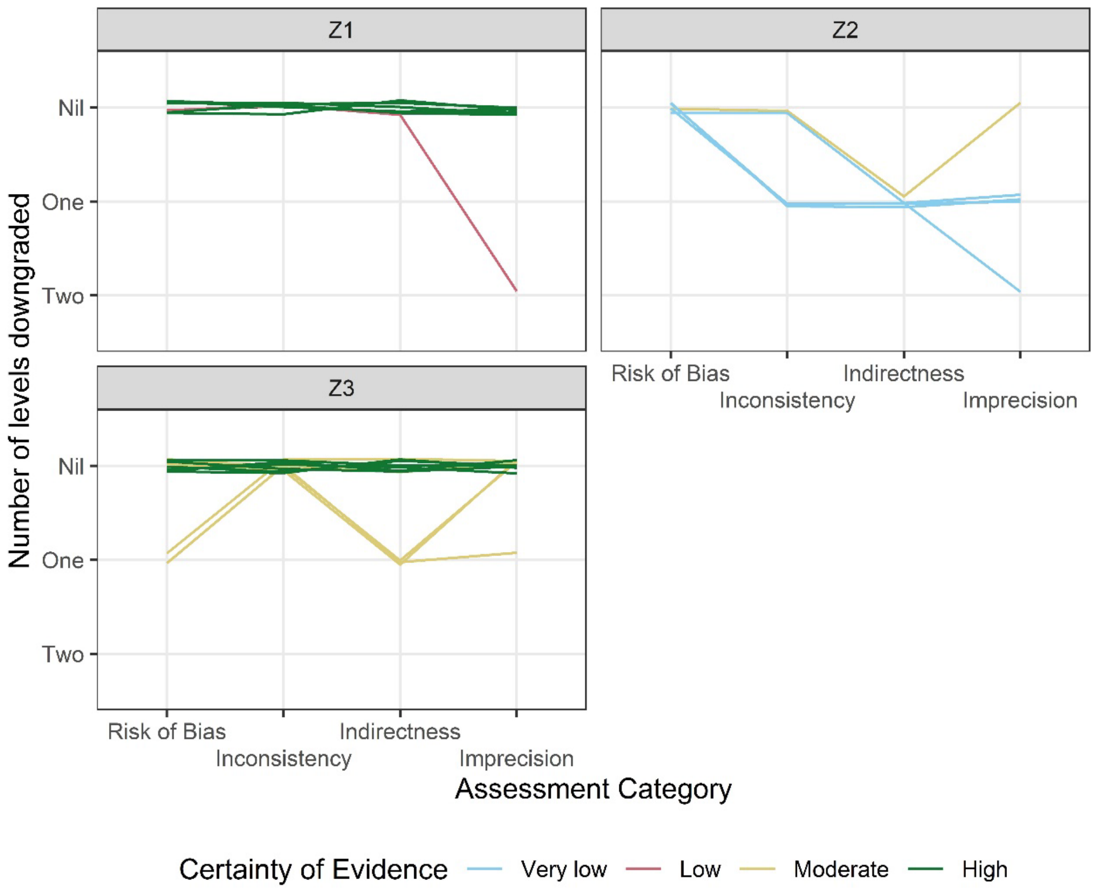


**Figure S6 Certainty of evidence classification for each outcome across each assessment category and research question for varicella zoster virus research questions. Outcomes classified High, Moderate, Low and Very Low have been downgraded zero, one, two and three or more levels, respectively.**
